# SARS-CoV-2 variants of concern, Gamma (P.1) and Delta (B.1.617), sensitive detection and quantification in wastewater employing direct RT-qPCR

**DOI:** 10.1101/2021.07.14.21260495

**Authors:** Karin Yaniv, Eden Ozer, Ariel Kushmaro

## Abstract

SARS-CoV-2 variants of concern present a worldwide threat. Demonstrating higher infection rate and durability to antibodies when compared to the original SARS-CoV-2 virus, the variants of concern are responsible for continuing global outbreaks. Prompt identification of the infecting SARS-CoV-2 variant is essential for pandemic assessment and containment. However, variant identification is mainly being performed using expensive, time-consuming next generation sequencing. Rapid identification methodology for variants of concern is of great need and various variant-specific assays are being developed. Amongst the variants of concern that have recently appeared, the Gamma variant (P.1, Brazilian) and Delta variant (B.1.617, Indian) are the most prominent. Here we describe the development of a sensitive RT-qPCR assay for the quick direct detection of the Gamma and Delta variants as part of a methodical characterization and detection in municipal wastewater.

## Introduction

Since declaring COVID-19 a world pandemic in March 2020, the disease continues to evolve and affect significant portion of the world population. Increasing circulation of the severe acute respiratory syndrome coronavirus 2 (SARS-CoV-2) virus within the world population, results in a continued natural processes of random mutations and evolution^1^. In some cases, a mutation will have an evolutionary advantage and create a new viral lineage that will overpower previous forms. First evidence for such an evolutionary event for the SARS-CoV-2 virus was in April 2020, when the D614G substitution enhanced infectivity and transmission. The D614G mutation lineage became the dominant SARS-CoV-2 variant form^2,3^. By late 2020, the term ’variants of concern’ emerged with regards to the high-numbered mutations in a single variant and the increased pathogenicity^4^.

Variants of concern are mainly characterized through mutations in their spike protein gene (S gene). Such mutations affect the receptor biding domain affinity^5^ and antibody neutralization efficiency^6^. Gamma (P.1, Brazilian) and Delta (B.1.617, Indian) variants of concern were first detected in January 2021 and December 2020 respectively, and by now circulate globally^7^. High transmission rates and lower vaccine efficacy towards these variants^8–10^ present a higher threat compared to the original virus. Therefore, quick detection of recognized variants of concern is vital for initiating response and proper policy for pandemic containment.

Currently, SARS-CoV-2 variant detection mainly relies on costly next generation sequencing, where results are obtained within 3-5 days^11,12^. With new variants of interest constantly emerging, sequencing-based approaches are important for their identification, however they present difficulties considering special instruments and resources, including data analysis skills required. To ease on detection a reverse transcriptase quantitative polymerase chain reaction (RT-qPCR)-based assay can provide rapid results for detection of known variants of concern detection. Such an assay can help assess a variant’s frequency within the population and contribute to policy guidelines. Indeed a number of publications targeting variants of concern detection using RT-qPCR^13–18^ as well as available commercial kits (TaqPath ThermoScientific, GT molecular, PerkinElmer, etc.), have emerged.

Urban morbidity monitoring through wastewater is a known practice^19^. Without the need for cooperation by individuals, it is possible to receive viable important information regarding the spread of a variant in population in a given area. Since erupting, the SARS-CoV-2 virus has been monitored in wastewater around the world^20–25^. In order to detect specific variants such as the Gamma and Delta variants of concern though, a direct and sensitive probe-based RT-qPCR assay with detection ability in wastewater samples is of great need and could help generate better health policies once employed in a given region. In this study, we present a sensitive RT-qPCR assay design for the direct detection of Gamma and Delta variants of concern and its employment in wastewater sampling.

## Methodology

### Primers and probes design

The original sequence of SARS-CoV-2 (NC_045512.2) was taken from NCBI database. Gamma variant (P.1, Brazilian, EPI_ISL_981709) and Delta variant (B.1.617, Indian, EPI_ISL_1704637) sequences were taken from GISAID database^7^. The probe design focused on the end of ORF8 and the beginning of N gene 28227-28286 bp location that includes the Gamma variant insertion, or S gene 21989-22083 bp location that includes the Delta variant deletion 157-158. All primers and probes were purchased through Integrated DNA Technologies (IDT). ZEN Quencher was added to the probes as a second, internal quencher in qPCR 5’-nuclease assay. Probes were assigned a 6-carboxy-fluorescein (FAM) fluorophore.

### Wastewater RNA extraction

Wastewater samples were collected in the city of Modiin, Israel for 24 hours composite sewage samples. Samples were taken from different manholes and immediately transferred to the lab under chilled conditions. Direct RNA was extracted twice according to Zymo Environmental Water RNA (Zymo Research R2042) manufacture protocol. For internal control, we added 10^5^ copies MS2 phage to the lysis buffer in each RNA extraction. RNA was eluted with 35 μL of RNase free water. The fresh wastewater samples were kept at 4°C until processed. RNA samples were kept at –80°C.

### RT-qPCR

RT-qPCR was executed as previously described^17^. Reaction final volume is 20 µL with primers concentration of 0.5 µM and probe concentration of 0.2 µM. The reaction contained 5 µL of RNA sample and ROX as a reference dye. Reaction steps was executed according to manufacture recommended protocol (One Step PrimeScript III RT-qPCR mix RR600 TAKARA, Japan) using Applied Biosystems, Thermo Scientific). Each RT-qPCR run included relevant quality controls, Non template control (NTC) and MS2 phage detection for wastewater RNA sample^26^.

### Calibration curves and limit of detection determination

Calibration curves were performed on a known-positive DNA gene block. Three different gene blocks were used; (1) containing SARS-CoV-2 S gene sequence as reported for Wuhan-Hu-1 (NC_045512.2):TACCCTGACAAAGTTTTCAGATCCTCAGTTTTACATTCAACTCAGGACTTGTTCTTACCTTTCTTTTCCAATGTTACTTGGTTCCATGCTATACATGTCTCTGGGACCAATGGTACTAAGAGGTTTGATAACCCTGTCCTACCATTTAATGATGGTGTTTATTTTGCTTCCACTGAGAAGTCTAACATAATAAGAGGCTGGATTTTTGGTACTACTTTAGATTCGAAGACCCAGTCCCTACTTATTGTTAATAACGCTACTAATGTTGTTATTAAAGTCTGTGAATTTCAATTTTGTAATGATCCATTTTTGGGTGTTTATTACCACAAAAACAACAAAAGTTGGATGGAAAGTGAGTTCAGAGTTTATTCTAGTGCGAATAATTGCACTTTTGAATATGTCTCTCAGCCTTTTCTTATGGACCTTGAAGGAAAACAGGGTAATTTCAAAAATCTTAGGGAATTTGTGTTTAAGAATATTGATGGTTATTTTAAAATATATTCTAAGCACACGCCTATTAATTTAGTGCGTGATCTCCCTCAGGGTTTTTCGGCTTTAGAACCATTGGTAGATTTGCCAATAGGTATTAACATCACTAGGTTTCAAACTTTACTTGCTTTACATAGAAGTTATTTGACTCCTGGTGATTCTTCTTCAGGTTGGACAGCTGGTGCTGCAGCTTATTATGTGGGTTATCTTCAACCTAGG; (2) containing the end of ORF8 until the beginning of N gene carried on a plasmid matching the reported 4 nucleotides insertion of the Gamma variant:TAATTGCCAGAAACCTAAATTGGGTAGTCTTGTAGTGCGTTGTTCGTTCTATGAAGACTTTTTAGAGTATCATGACGTTCGTGTTGTTTTAGATTTCATCTAAACGAACAAACAAACTAAAATGTCTGATAATGGACCCCAAAATCAGCGAAATGCACCCCGCATTACGTTTGGTGGACCCTCAGATTCAACTGGCAGTAACCAGAATGGAGAACGCAGTGGGGC; (3) containing S gene sequence matching the reported 157-158 deletion of the Delta variant: TTGTTATTAAAGTCTGTGAATTTCAATTTTGTAATGATCCATTTTTGGATGTTTATTACCACAAAAACAACAAAAGTTGGATGGAAAGTGGAGTTTATTCTAGTGCGAATAATTGCACTTTTGAATATGTCTCTCAGCCTTTTCTTATGGACCTTGAAGGAAAACAGGGTAATTTCAAAAATCTTA GGG. Calibration of P.1 probe was performed using the second gene block, while calibration of SΔ157 probe was performed with the third gene block. Negative control for the P.1 probe was performed using Wastewater samples confirmed as positive for the original variant or Alpha variant of concern, and negative control for the SΔ157 probe was performed using the first gene block. Serial dilutions for the relevant gene block were prepared based on copy number calculations. The resulting Ct values were plotted against the log copy number of the gene block template. Each concentration was examined by six repetitions and a standard deviation was calculated. Linear regression was performed between the log copy number and the Ct values from the RT-qPCR results.

### Complex matrix detection

In order to validate the RT-qPCR designed assay’s sensitivity in a wastewater matrix, RNA extracted from pre-determined negative wastewater sample for SARS-CoV-2 was examined using standard CDC’s detection sets. The negative RNA sample was supplemented with known concentrations of a desired gene block (10^0^/10^1^/10^2^). All experiments executed with the matrix pre-determined as negative for additional verification of the sample as negative. Eight repetitions were performed for each viral concentration or control. Ct results were plotted to represent the new probes limit of detection in a complex environment.

## Results and Discussion

Following the Alpha variant B.1.1.7 (British) eruption and containment in Israel, the Gamma and Delta variants were believed to be the most urgent variants in need for rapid detection. Previously we developed direct RT-qPCR detection assays for Alpha variant (B.1.17, British) and Beta variant (B.1.315, South Africa)^17^. During the new outbreak we further developed a direct RT-qPCR detection assays for Gamma variant (P.1, Brazilian) and Delta variant (B.1.617, Indian). Our design for detection of these two variants (Fig. 1) is based on the differences in gene sequence from the original SARS-CoV-2 sequence (NC_045512.2). The Gamma variant contains an insertion in the ORF8 region, and the Delta variant S gene contains a deletion known as Δ157-158. Accordingly, our designed primers-probe sets for variants detection focused on these regions and is presented in Figure 1.

**Figure 1.**
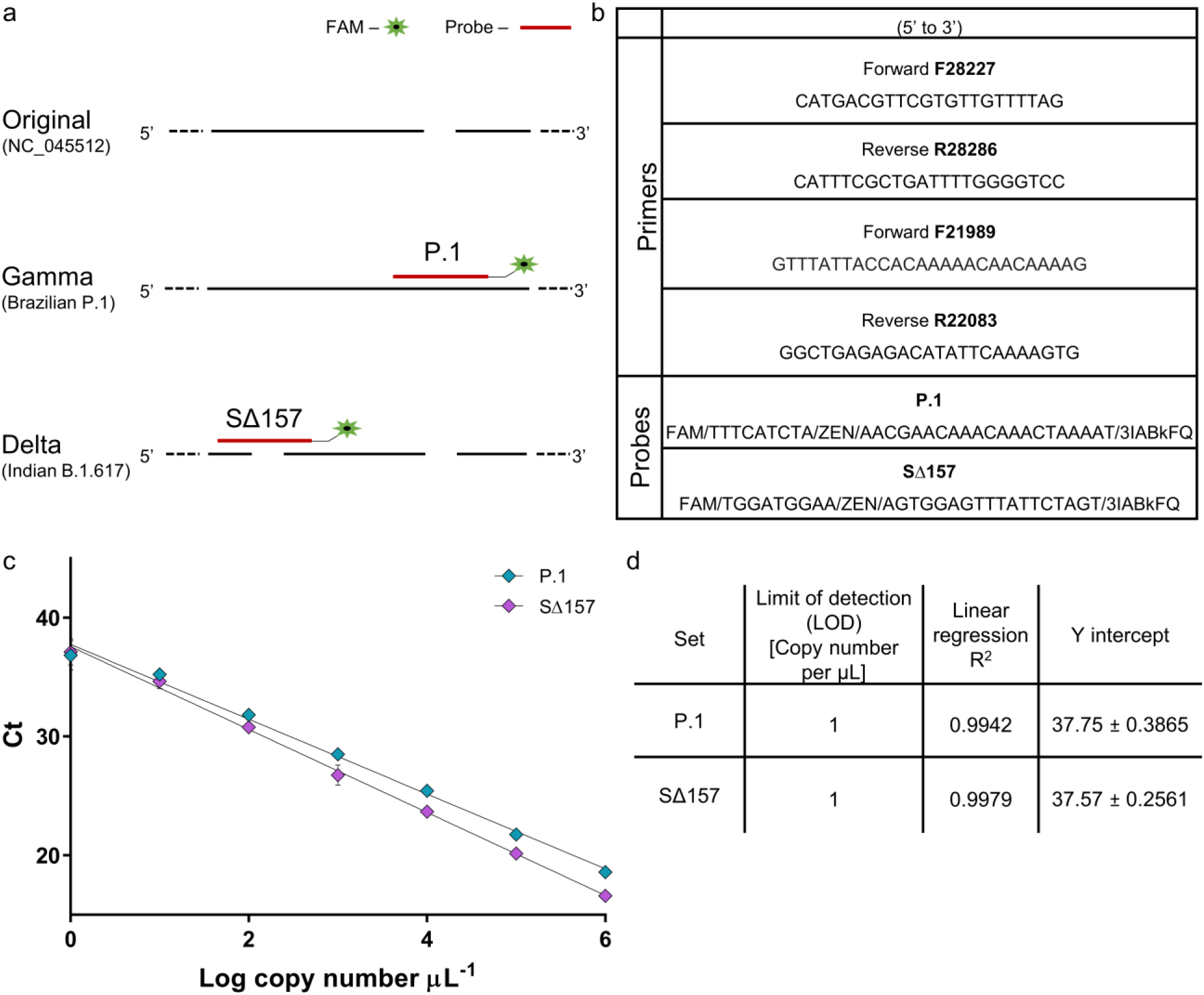
RT-q-PCR primers-probe sets design for Gamma and Delta variants detection. (a) Designed detection sets for differentiation and identification of SARS-CoV-2 Gamma variant (P.1) and Delta variant (B.1.617). Design was based on sequence changes between the original SARS-CoV-2 virus and variants of concern, with insertion after ORF8 for Gamma variant and deletion in the S gene of the Delta variant. (b) List of primers and probes sequences. (c) Calibration curves for primers-probe sets developed for variants of concern detection. Resulted Ct value plotted against the tested Log copy number. Error bars present standard deviation for ten replicates. Positive DNA template for SΔ157 set, corresponded to Δ156-157 deletion site in B.1.617 S gene. Positive DNA template for P.1 set, corresponded to the DNA segment from the end of ORF8 and the beginning of N gene including the 4 nucleotides insertion. (d) List of Limit of detection, Linear regression R^2^, and Y intercept values extracted from the devised calibration curves.

For Gamma variant detection, the designed forward primer is located at the end of ORF8 gene and the reverse primer is located at the beginning of the nucleocapsid gene (N gene), meaning 28227-28286 bp of the original sequence. Within this range, the original SARS-CoV-2 and Gamma variant sequences are completely identical, apart from a 4 nucleotides insertion in the Gamma lineage (Fig. 1a). Using a detection set comprised of two primers meant to amplify the target region, a single probe (P.1 probe) was designed for the detection of the Gamma variant only (corresponding to the insertion). For Delta variant detection, the designated detection region was chosen from within the S gene. Focusing on S gene’s 21989-22083 bp of the original sequence, the original SARS-CoV-2 sequence is identical to the Delta variant sequence with the exception of a 6 nucleotides deletion in the Delta lineage (Fig. 1a). Using a detection set comprised of two primers meant to amplify the target region, a single probe (SΔ157 probe) was designed for the detection of the Delta variant only (corresponding to the deletion).

To ensure functionality, the described sets of primers and probes underwent characterization. Initially, a calibration curve was generated for the two primers-probe sets, using dsDNA as a template. A detection range of between 10^6^ copies and 10^0^ copies per µL was tested for each set (Fig. 1c). Linear regression performed for the two probes demonstrated strong correlation. A limit of detection (LOD) could be determined for each primers-probe set and was identified as 10^0^ copies per µL for the two sets (Fig. 1d), meaning highly sensitive detection. Specificity of the described sets was examined with either dsDNA fragments from the original SARS-CoV-2, relevant to the assigned detection regions, or using wastewater found positive for the original SARS-CoV-2 and the Alpha variant. Both sets did not manifest a signal when tested with such negative controls and were therefore found to be highly specific.

Primers-probe sets were further characterized within a more intricate matrix. Wastewater from Binyamina, Israel served as a more complex environment for the RT-qPCR assay, as sewage samples contain a variety of materials and organisms. Wastewater samples were pre-determined as negative for SARS-CoV-2 using N gene detection. After confirming that the wastewater samples were negative, relevant dsDNA template copies were added exogenously at known concentrations (Fig. 2). These samples were then used for detection by the examined primers-probe sets. As can be seen in Figure 2, despite the wastewater matrix, P.1 and SΔ157 probes maintained high functionality with a LOD of 10^0^ copies per µL, proving their sensitivity and stability.

**Figure 2.**
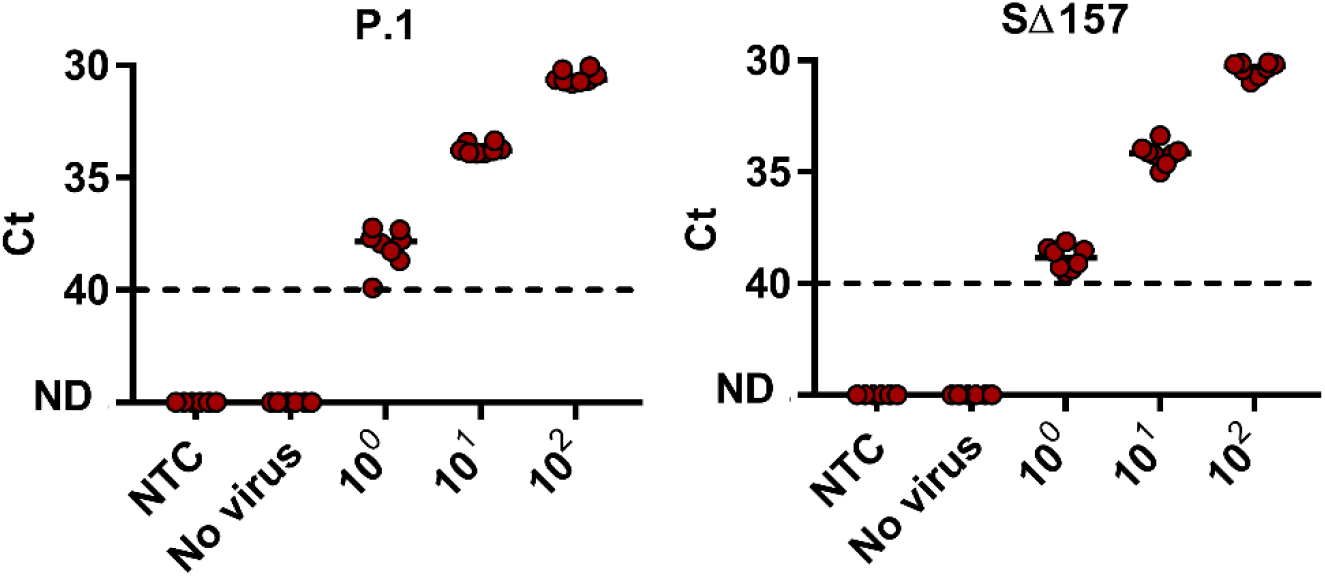
Lower detection limit of P.1 and SΔ157 primers–probe sets in wastewater matrix. RNA extracted from negative detection wastewater sample (No virus) spiked with known concentrations of positive template corresponding to each probe accordingly (10^0^–10^2^ S gene template copies per μL) and Non-Template Control (NTC, water). Positive DNA template for P.1 set, corresponded to the DNA segment from the end of ORF8 and the beginning of N gene including the 4 nucleotides insertion in the Gamma variant. Positive DNA template for SΔ157 set, corresponded to Δ156-157 deletion site in the Delta variant. ND - not detected. Solid lines indicate the median and dashed lines indicate the detection limit as decided by clinical guidelines.

After fully characterizing the developed primers-probe sets, revealing their specificity, sensitivity and durability, it was possible to employ them on recently collected wastewater samples. Until recently, the Alpha variant was most dominant in Israel, however a noticeable decrease was observed in the past months. In the past month (May-June 2021), reports indicated that the Delta variant had reached Israel and is responsible for the majority of new morbidity cases. Knowing this, samples from different locations at the city of Modi’in were collected and tested for general SARS-CoV-2 presence, using N gene detection, and Alpha and Delta variants presence, using relevant S gene detection (Table 1).

**Table 1:**
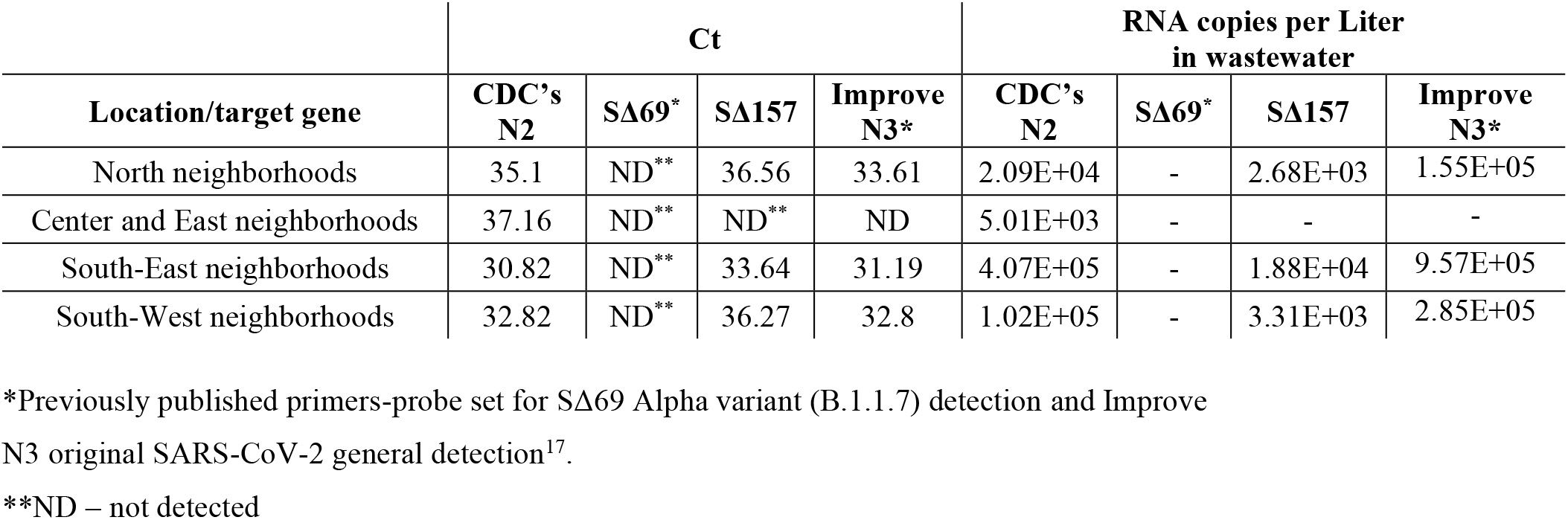
Modi’in city manholes detection for SARS-CoV-2 N gene (CDC’s N2, Improved N3), Alpha variant S gene (SΔ69) and Delta variant S gene (SΔ157)

As can be seen in Table 1, all of the tested samples were positive to SARS-CoV-2 presence through N gene detection. However, none of the samples resulted in a positive detection for the Alpha variant, corresponding to the disappearance of this variant in clinical tests as reported by health officials. Furthermore, all samples, apart from one, resulted in a positive signal for the Delta variant using the designed primers-probe set described here. These field results indicated the new probe sets ability to correctly detect and quantify the Delta variant in a sensitive manner, even within a complex environment, and provide an assessment for variant of concern geographical spread in the population. Considering the importance of wastewater monitoring, as well as the possibility of clinical employment, we hope the developed sets can serve for variants of concern detection.

## Data Availability

All data needed to evaluate the conclusions in the paper are present in the paper

## Author Contributions

K.Y. and E.O. share equal contribution to this manuscript. K.Y. designed sequence, conceived, performed and analyzed experiments and authored this manuscript. E.O. designed sequence, conceived and analyzed experiments and authored this manuscript. A.K. conceived experiments, supervised, provided research facilities and edited the manuscript.

## Acknowledgement

We thank KANDO and the Israeli Ministry of Health for providing us with the sewage samples. We gratefully acknowledge GISAID database for access to SARS-CoV-2 variants sequences. We gratefully acknowledge Esti Kramarsky-Winter assistance for comments and scientific editing of the manuscript.

